# TREATMENT COSTS FOR COVID-19 PATIENTS IN A TERTIARY HOSPITAL FROM SERBIA

**DOI:** 10.1101/2021.11.30.21267085

**Authors:** Predrag S. Sazdanovic, Slobodan Milisavljevic, Dragan R. Milovanovic, Slobodan M. Jankovic, Dejan Baskic, Dragana Ignjatovic Ristic, Dejana Ruzic Zecevic, Aleksandra Tomic Lucic, Natasa Djordjevic, Danijela Jovanovic, Andjelka Stojkovic, Tatjana Lazarevic, Milica Begovic Cvetkovic, Marina J. Kostic

## Abstract

**Introduction:** Aim of our study was to identify total costs of COVID-19 inpatients treatment in an upper-middle income country from Southeast Europe.

**Methods:** This retrospective, observational cost of illness study was performed from National Health Insurance Fund perspective and included a cohort of 118 males and 78 females admitted to COVID-19 ward units of a tertiary center, during the first wave of epidemics.

**Results:** The median of total costs in the non-survivors’ subgroup (n=43) was 3279.16 Euro (4023.34, 355.20, 9909.61) which is higher than in the survivors (n=153) subgroup 747.10 Euro (1088.21, 46.71, 3265.91). The odds ratio of Charlson Comorbidity Index total score and every 100-Euros increase of patient’s total hospital treatment costs for fatal outcome were 1.804 (95% confidence interval 1.408-2.311, p<0.001) and 1.050 (1.029-1.072, p<0.001), respectively.

**Conclusions:** Direct medical treatment costs for COVID-19 inpatients represent significant economic burden. The link between increased costs and unfavorable final outcome should be further explored.

## INTRODUCTION

The COVID-19 pandemic has unprecedented socio-economic implications worldwide and the evaluators projected billions of direct and indirect financial spending and losses for both individual countries and global workforce and economy (1, 2). Healthcare systems are among many sectors which experienced losses of revenues and incomes caused by disturbances of managing their regular operations. Especially hospitals are very vulnerable to the economic constraints of current pandemic, particularly academic centers (3). They are obliged to implement a multitude of preventive measures, perform various diagnostic procedures and provide a range of therapeutic approaches within rapidly changing and complicating institutional and societal environment (4). Unsurprisingly, the investigators from some developed countries reported that the spreading of the disease correlated with negative trends such as decline of elective hospital services, decrease of overall hospitalization rate, reduction of claimed hospital charges and lowering of their financial gains (5, 6).

The substantial impact of COVID-19 on economic performances of healthcare systems needs to be analyzed more comprehensively. However, there are a few published studies based on or modelled for hospital data. Some of the examples are reports from Saudi Arabia and South Africa. They estimated direct medical expenses for hospital treatment of COVID-19 patients, showing that costs per patient almost doubled in comparison to period before the pandemy (7, 8). Additionally, data from hospitals in the United States confirmed that intensive management of COVID-19 inpatients (e.g. critical unit care, mechanical ventilation) generated high financial burden, also indicating large financial gap between charges and cost claims per treated subject (9). Current situation in majority of other countries remains insufficiently documented for the time being.

The health care analysts, planners and managers could reasonably assume that COVID-19 pandemy seriously undermined economic sustainability of modern hospitals. However, it is necessary to bring additional evidence about main cost drivers and their mitigating factors, comparing a variety institutional settings from different societal, cultural and economic environments. Therefore, the aim of our study was to perform detailed analysis of COVID-19 inpatients treatment costs in an upper-middle income country from Southeast Europe.

## PATIENTS AND METHODS

The research was designed as a cost-study. The sample was based on observational approach and it included a cohort of 196 adult patients (males and females, 18 years and older) admitted to COVID-19 ward units of University Clinical Center “Kragujevac”, Kragujevac, Serbia, between March 14th and April 26th, 2020, during the first wave of epidemics. The sample represents approximately a three-quarter of all patients which had been admitted to the hospital COVID-19 units during that period. The study group was divided to case (the non-survivors) and control patients (the survivors). We extracted data retrospectively from the patients’ electronic medical records. The general design of our trial was based on previously published clinical studies with similar, observational design which included COVID-19 inpatients (10, 11). The study was approved by the Institutional Ethics Committee and conducted according to principles of Declaration of Helsinki (Decision of the Ethics Committee number 01/20-407, dated April 3^rd^ 2020).

The patients’ demographic and clinical data during the hospital treatment period were transformed to study variables, selected from a set of recommendations for building case report forms for patients with COVID-19 enrolled to clinical trials (12). COVID-19 severity and clinical improvement were assessed according to the recommendations of World Health Organization (13, 14). We used the International Statistical Classification of Diseases and Related Health Problems, 10th Revision for classifying comorbid disorders (15). Charlson Comorbidity Index (CCI) total score showed patient’s pre-existing illness burden on admission together with his or her 10-year probability of survival (16).

The cost analysis included direct medical cost for patient treatment during hospital stay. We used the perspective of National Health Insurance Fund with the official price tariffs for hospital health care services, reimbursed prescription drugs and licensed medical devices (so called “hospital electronic bill”) which were established for fiscal year 2020. (www.rfzo.rs/index.php/davaocizdrusluga/efaktura). The original prices in Serbian Dinars (RSD) had been converted to Euros (EUR) with the first exchanged rate of National Bank of Serbia for 2020, released on January the 3^rd^ (1 EUR: 117.5967 RSD) (www.nbs.rs). The calculated costs are compared with average wages in Serbia, according to the official data issued by national Statistical Office (www.stat.gov.rs/sr-Latn/oblasti/trziste-rada/zarade).

There are two cost-drivers of medical treatment in our hospital - admission units and hospital wards. In general, the patient’s stay in admission units was very short, it did not go beyond one day, so for these units only total costs were showed. The costs for ward management of the patients included medical services (e.g. physician examinations, nurse care, clinical biochemistry and pathology analyses, radiological examinations), drugs and medical devices (excluding biochemical reagents or diverse laboratory consumables as they had been billed cumulatively, every month, on hospital level).

The analysis of collected data included descriptive statistics (measures of central tendency and variations) and comparison between the study groups depending on the type and data distribution (t-test, Mann-Whitney test, χ^2^-test or Fisher’s exact test, Pearson’s and Spearman’s correlation). Binary logistic regression included multivariable approach, too; however, we imputed very limited set of study variables, taking into account methodological and sample-size limitation, in order to avoid flawed outputs (e.g. over fitting). Receiver-operating characteristic curve (ROC) estimation was a tool for analysis of predictive performance of total costs for patient final outcomes. For all calculations the probability of null hypothesis ≤0.05 was considered to be statistically significant.

## RESULTS

Majority of the study population were men, being in the middle of the sixth decade of life. There were 150 (76.5%) subjects with positive test of polymerase chain reaction (PCR) for SARS-CoV-2 and the others were hospitalized based on sound clinical and/or epidemiological features strongly suggestive for COVID-19 disease. Main demographic and clinical characteristics of the study subjects are presented in the Table 1. Additional co morbid conditions at hospital admission were moderate-to-severe chronic kidney disease and connective tissue disease within fatal outcome group in 6 and 1 patients, respectively as well as leukemia in 1 patient who survived; no formal diagnoses of peripheral vascular disease, dementia, liver disease, hemiplegia, lymphoma and acquired immunodeficiency syndrome had been documented in the patients’ medical files. Information about symptom onset were available for 158 (80.6%) patients and for that subgroup COVID-19 medical treatment lasted about three weeks. The case fatality rate was 21.9%, as 43 study subjects died during the hospital treatment.

**Table 1.** Demographic and main clinical characteristics of study patients.

| Variable | All patients<br>(n=196) | Survivors<br>(n=153) | Non-survivors<br>(n=43) | Statistics<br>(test; p) |
| --- | --- | --- | --- | --- |
| Age (years) | 58.4±15.3<br>(60, 21, 19-88) | 55.6±14.9<br>(58, 22, 19-88) | 68.3±12.4<br>(70, 20, 45-88) | t=5.1;<br><0.001* |
| Gender (male) | 118 (60.2) | 92 (60.1) | 26 (60.5) | $\chi^2$<0.1;<br>0.968 |
| Hypertension | 87 (44.4) | 65 (42.5) | 22 (51.2) | $\chi^2$=1.0;<br>0.312 |
| Myocardial infarction<br>history | 12 (6.1) | 9 (5.9) | 3 (7.0) | $\chi^2$<0.1;<br>0.728 |
| Chronic heart failure | 8 (4.1) | 3 (2.0) | 5 (11.6) | $\chi^2$=8.1;<br>0.014* |
| Cerebrovascular accident or<br>TIA history | 3 (1.5) | 1 (0.7) | 2 (4.7) | $\chi^2$=3.6;<br>0.122 |
| Chronic obstructive<br>pulmonary disease | 28 (14.3) | 13 (8.5) | 15 (34.9) | $\chi^2$=19.1;<br><0.001* |
| Peptic ulcer disease | 5 (2.6) | 3 (2.0) | 2 (4.7) | $\chi^2$=1.0;<br>0.302 |
| Diabetes mellitus,<br>uncomplicated | 16 (8.2) | 12 (7.8) | 4 (9.3) | $\chi^2$=0.1;<br>0.756 |
| Diabetes mellitus,<br>complicated | 19 (9.7) | 12 (7.8) | 7 (16.3) | $\chi^2$=0.1;<br>0.756 |
| Solid tumor | 3 (1.5) | 2 (1.3) | 1 (2.3) | $\chi^2$=2.7;<br>0.140 |
| CCI, score | 2.3±1.9<br>(2, 3, 0-8) | 1.8±1.7<br>(2, 3, 0-8) | 3.8±1.8<br>(4, 3, 1-8) | z=-5.9;<br><0.001* |
| CCI, survival | 75.9±29.2<br>(90, 43, 0-98) | 82.6±23.6<br>(90, 21, 0-98) | 51.9±34.3<br>(53, 69, 0-96) |  |
| COVID-19 disease duration<br>/ hospital treatment (days)** | 23.4±8.1 (23, 9, 4-53; 158) | 23.8±8.1 (23, 10,<br>4-53; 128) | 21.5±8.1 (21,<br>13, 7-38; 30) | t=1.4;<br>0.161 |
| Hospital stay (days)** | 15.2±7.0 (15, 7, 1-39) | 15.8±6.6 (15, 7,<br>1-39) | 13.4±8.1 (12,<br>11, 1-30) | z=-2.3;<br><0.024* |
| Pneumonia | 160 (81.6) | 117 (76.5) | 43 (100) | $\chi^2$=12.4;<br><0.001* |
| Sepsis | 64 (32.7) | 21 (13.7) | 43 (100) | $\chi^2$=113.6;<br><0.001 |
| Septic shock | 26 (13.3) | 2 (1.3) | 24 (55.8) | $\chi^2$=86.7;<br><0.001* |
| Acute coronary syndrome | 8 (4.1) | 3 (2.0) | 5 (11.6) | $\chi^2$=8.1;<br>0.014* |
| High-flow oxygen | 15 (7.7) | 11 (7.2) | 4 (9.3) | $\chi^2$=14.5;<br>0.001* |
| Non-invasive ventilation | 2 (1.0) | 1 (0.7) | 1 (2.3) |  |
| Mechanical ventilation | 45 (23.0) | 9 (5.9) | 36 (83.7) |  |
| Duration of mechanical<br>ventilation (h) | 247.4±161.8 (209,<br>246, 18-735; 40) | 242.6±136.6<br>(202, 231, 72-500; 13) | 249.7±175.1<br>(216, 269, 18-735; 27) | t=0.1,<br>0.899 |

the numbers represent the mean ± standard deviation (median, interquartile range, minimal, maximal; number of patients) or number (percent) of patients, as appropriate; z - Mann-Whitney test; t - Student's t-test; $\chi^2$ - Fisher's exact or $\chi^2$ test; p - probability for difference between the groups; \*-significant difference; CCI- Charlson Comorbidity Index, estimated 10-year survival; \*\*data for prehospital symptoms and treatments were missing for 39 patients and hospital stay was identified for 158 subjects

Distribution of COVID-19 disease severity in study cohort was as following: 5 (2.6%) asymptomatic patients, 31 (15.8%) subjects with mild disease, 64 (32.7%) patients with moderate disease (pneumonia), 32 (16.3%) patients with severe disease (severe pneumonia), and 64 (32.7%) people with critical disease. All patients in the non-survivor group had developed critical illness before death. Distribution of the clinical improvement score within the patients who survived was as following: score 3 in 85 (55.6%), 4 in 47 (30.7%), 5 in 12 (7.8%), 6 in 7 (4.6%) and 7 in 2 (1.3%) patients, respectively. The clinical improvement score of 8 was finally assigned to all patients who succumbed the disease. Beside COVID-19 complication noted within the table acute respiratory distress syndrome was documented in 7 (16.3) patients who died but not among the survivors; delirium was diagnosed formally in 5 (3.3) patients, all of which recovered. Very short hospital stay (<5 days) was noted for 8 (4.1%) patients, but since costs for their treatment were still substantial they were included in the final analysis set.

Direct medical cost of hospital care per a patient in COVID-19 units was substantial (Table 2). The median total cost for the whole study cohort, compared to the national average monthly salary expressed as all amount of earnings (brutto) and wages without taxes and obligatory insurance expenses (netto), was higher by 39.6% and 92.9%, respectively. In addition, the costs were significantly higher for subjects of the non-survivor group in comparison with the patients who survived (p<0.001), except for cost at hospital admission units.

**Table 2.** Direct medical cost of hospital care (EUR) in COVID-19 units, per a patient, for total duration of hospital stay.

| Costs | All patients<br>(n=196) | Survivors<br>(n=153) | Non-survivors (n=43) | Statistics<br>(test; p) |
| --- | --- | --- | --- | --- |
| Admission<br>units | 4.62±14.64 (2.41, 0, 0-138.85) | 4.18±13.41<br>(2.41, 0, 0-138.85) | 6.20±18.45<br>(2.41, 3.94, 0-105.68) | z=-0.0;<br>0.990 |
| Ward unit<br>services | 1023±878.34 (665.77,<br>823.00, 0-5685.00) | 827.08±732.29<br>(569.04, 481.71, 0-5685.00) | 1722.68±1000.06<br>(1514.10, 1457.86,<br>246.83-3929.43) | z=-5.9;<br><0.001* |
| Ward unit<br>drugs | 599.80±920.61<br>(201.70, 783.22, 0-6227.15) | 377.38±685.30<br>(125.93, 467.24, 0-6227.15) | 1391.17±1189.07<br>(1030.09, 1702.60,<br>15.73-5466.84) | z=-6.6;<br><0.001* |
| Ward unit<br>medical<br>devices | 248.62±432.73 (69.30,<br>271.04, 0.40-3298.01) | 146.19±266.58 (40.28,<br>124.39, 0.40-1400.30) | 613.07±661.80<br>(400.52, 687.37,<br>13.86-3298.01) | z=-6.9;<br><0.001* |
| Total | 1876.60±2083.00<br>(983.04, 1818.83,<br>146.71-13265.91) | 1354.84±1608.03<br>(747.10, 1088.21,<br>146.71-13265.91) | 3733.11±2503.28<br>(3279.16, 4023.34,<br>355.20-9909.61) | z=-6.5;<br><0.001* |

the numbers represent the mean ± standard deviation (median, interquartile range, minimal-maximal), as appropriate; z - Mann-Whitney test; \*-significant difference

The median of patients’ costs for ward health-care services, ward drugs, ward medical devices and total costs were about 2.7, 2.2, 10 and 4.4 times higher, respectively, for the group with fatal outcome than for the group of patient who recovered the diseases, the difference being highly statistically significant. Logarithmic base-10 transformed total cost data (done to provide normal data distribution) were positively correlated with patients age (Pearson’s r=0.381, <0.001), Charlson Comorbidity Index total score (Spearman’s rho=0.465, p<0.001) and duration of treatment within the hospital (Pearson’s r=0.585, <0.001).

In addition, every one-point increase of patient’s CCI total score and every 100-Euros increase of patient’s total hospital treatment costs elevated significantly the odds for fatal outcome by 80.4% (odds ratio 1.804, 95% confidence interval 1.408-2.311, p<0.001) and 5.0% (odds ratio 1.050, 95% confidence interval 1.029-1.072, p<0.001), respectively (the model with two variables). ROC curve for prediction of fatal outcome based on the total cost had area under the curve (AUC) of 0.825 (95% confidence interval 0.752-0.897, p<0.001) (Fig. 1). The cut-off value (calculated by the Youden’s rule) of 1930.61 Euro of the total costs had 74.4% sensitivity and 81.7% specificity for prediction of patients’ dying from COVID-19 disease during hospital treatment.

**Figure 1.**
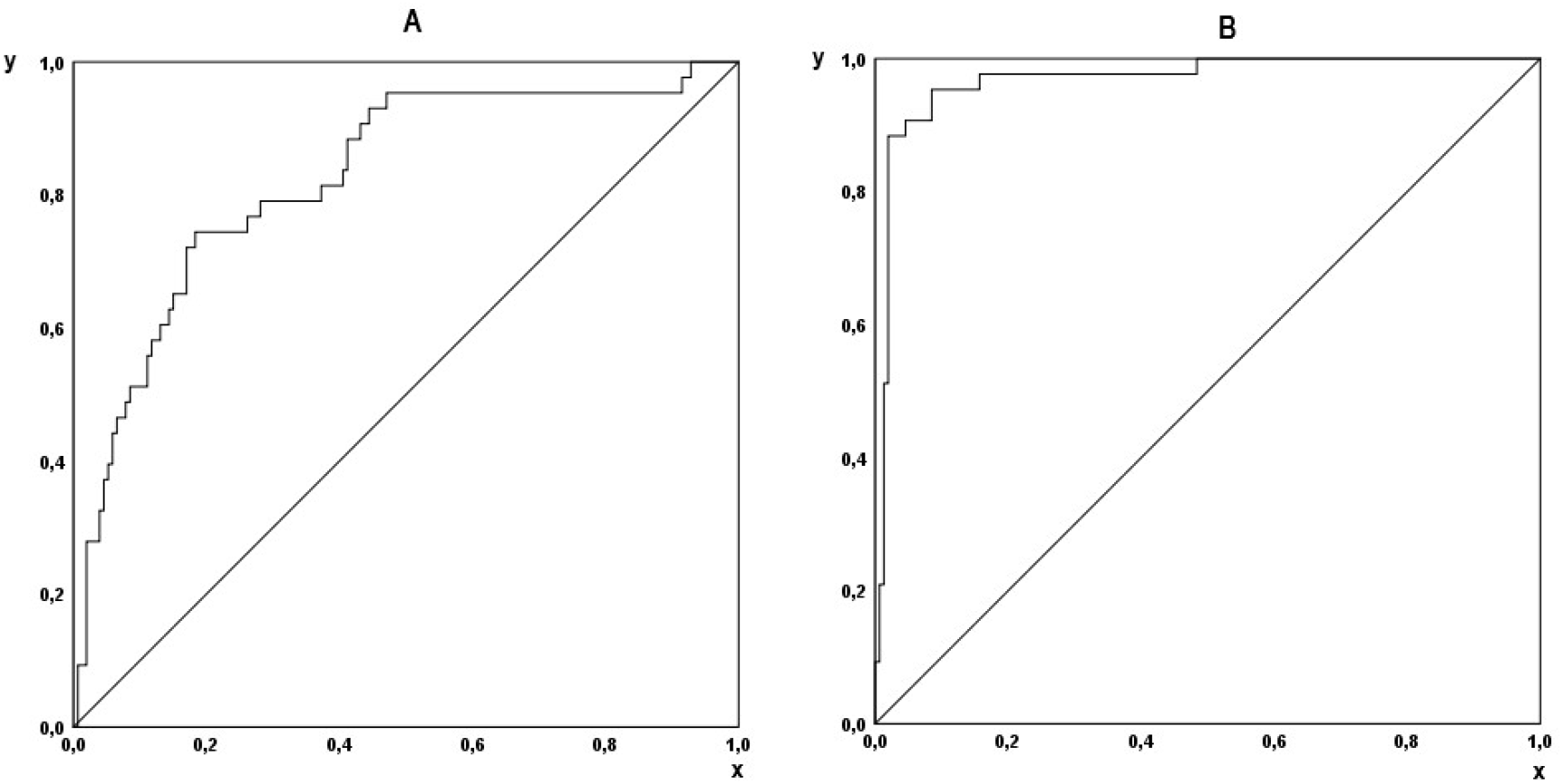
Receiver operating curve (ROC) for prediction of fatal outcome based on the total direct medical costs per patient (panel A) and direct medical costs, per patient per day of hospital treatment (panel B); y axes: sensitivity, x axes: 1 - specificity.

The additional cost analysis included standardized values - costs per patient per day of hospital treatment. Taking into account these data the median of patients’ costs for ward health-care services, ward drugs, ward medical devices and total costs were about 3.0, 9.1, 10.9 and 4.6 times higher, respectively, for the group with fatal outcome than for the group of patient who recovered the diseases, the difference being highly statistical significant (Table 3). Logarithmic base-10 transformed total cost per a day were positively correlated with patients age (Pearson’s r=0.330, p<0.001) and CCI total score (Spearman’s rho=0.420, p<0.001), too.

**Table 3.** Direct medical cost of hospital care (EUR) in COVID-19 units, per a patient for standardized cost data (per day of hospital stay).

| Costs | All patients<br>(n=196) | Survivors<br>(n=153) | Non-survivors (n=43) | Statistics<br>(test; p) |
| --- | --- | --- | --- | --- |
| Admission<br>units | 0.33±0.76 (0.16, 0.13,<br>0-6.61) | 0.31±0.73 (0.16,<br>0.11, 0-6.61) | 0.40±0.87 (0.17, 0.36,<br>0-5.03) | z=-0.2;<br>0.817 |
| Ward unit<br>services | 71.27±51.85 (57.73,<br>58.35, 0-360.66) | 52.13±34.41 (44.31,<br>36.10, 0-360.66) | 139.36±45.97<br>(133.91, 30.88, 58.56-304.84) | z=-9.5;<br><0.001* |
| Ward unit<br>drugs | 36.12±41.59 (19.90,<br>50.60, 0-182.23) | 20.05±24.28 (9.98,<br>29.03, 0-159.67) | 93.29±40.16 (91.00,<br>57.18, 2.62-182.23) | z=-8.8;<br><0.001* |
| Ward unit<br>medical<br>devices | 16.99±29.54 (5.74,<br>18.22, 0.04-194.00) | 8.36±15.28 (3.21,<br>9.22, 0.04-144.25) | 47.70±44.41 (35.07,<br>35.14, 2.31-194.00) | z=-8.4;<br><0.001* |
| Total | 124.71±106.01 (87.66,<br>122.94, 23.54-512.93) | 80.85±59.18 (60.33,<br>69.19, 23.54-409.60) | 280.75±87.00<br>(275.90, 96.74, 64.15-512.93) | z=-9.4;<br><0.001* |

the numbers represent the mean ± standard deviation (median, interquartile range, minimal-maximal), as appropriate; z - Mann-Whitney test; \*-significant difference;

In addition, every one-point increase of patient’s CCI total score and every 10-Euros increase of patient’s total hospital treatment costs per a day of hospital treatment elevated significantly the odds for fatal outcome by 2.5 times (odds ratio 2.517, 95% confidence interval 1.575-4.022, p<0.001) and 36.0% (odds ratio 1.360, 95% confidence interval 1.226-4.509, p<0.001), respectively. ROC for prediction of fatal outcome based on the total cost per a day had AUC of 0.968 (95% confidence interval 0.940-0.996, p<0.001) (figure 2). The cut-off value (according to the Youden’s rule) of 156.46 Euro of the total costs per a day had 95.3% sensitivity and 91.5% specificity for prediction of patients’ dying from COVID-19 disease during hospital treatment.

## DISCUSSION

Our study showed that the direct medical cost for the treatment of COVID-19 inpatient within the tertiary hospital settings were substantial and that they were significantly higher for patients with fatal outcome. The cost for medical services during the ward stay dominated among the other cost types, which included prescription drugs, medical devices and ambulatory admission care. The comorbidity burden at admission, together with patient’s age, duration of hospital treatment and the organ support measures, were the main cost drivers. Moreover, patient’s comorbidity and total costs were independent predictors of the fatal outcome. We quantified the magnitude and the variability of the influence of these factors on total direct medical costs and proposed the cost to be a marker of poor prognosis with a fair diagnostic performance. Taking into account the paucity of similar data we consider our study significant and original contribution to the topic.

To compare of our data with the results of other studies one has to take into account patients’ characteristics and socio-economic and cultural differences among countries. It seems that patients in our study more frequently had severe forms of chronic diseases at admission and severe type of COVID-19 with longer hospital stay (∼2-3 times), more mechanical ventilation treatments (up to the three quarter) and higher case fatality rate (up to the two third) than those reported from Saudi Arabia and United States (7, 9). Some differences in patients’ age and gender also exist between the three studies. Despite of these facts, the median of total costs in our study were several times lower than in other two countries and similar difference remains when the comparison included the results of the modeling study from South Africa, too (8).

We could look at the data of a standardized economic indicator such as gross domestic products at purchasing power parity per capita (GDP PPP) for these countries, trying to mitigate the influence of societal and health care system differences on the study data extrapolations (17). Such exercise revealed ∼3 times less GDP PPP-to-cost ratio based on our study data (representing an upper middle income society-Serbia) than the respective ratios based on data from two above-mentioned studies from high-income countries – United States and Saudi Arabia (18). Our results put the cost data into the context of the average national workers’ earrings per month, too. These relations could be used as an additional tool for appropriate economic comparisons between existing and future similar research in the field around the globe.

Several limitations of our study exist such as, the single-center scope, the shortcomings of observational design with retrospective data collection, and the absence of adjustment of study outcomes for some important factors (e.g. COVID-19 complications, prescription drugs, care within intensive care units). We analyzed direct costs for patients’ medical treatment only, and we did not take into consideration the effects of indirect factors on overall cost burden for hospital economic performances, such as implementation of infection control measures or changes of working productivity of hospital personnel due to the psychological stress (19, 20). In general, the logistic reasons (e.g. technical constraints of hospital database, managerial obstacles) precluded more comprehensive methodological approach in our study.

On the other side, we believe that health economic researchers have to account one circumstance of particular interest - a study time-frame within the dynamics of epidemic course. The evidences are accumulating that changing of SARS-CoV-2 biology and pattern of COVID-19 disease, as well as improvement of preventive, diagnostic and treatment measures, significantly affect many individual and societal outcomes related to the pandemics. For example, the in-hospital case-fatality rate from COVID-19 in United States decreased from 22.1% at the beginnings of epidemics (similar to our results), to 6.5% after several months (21). In the meantime, treatment guidelines incorporated new evidence, advising the clear shift from diversity of empiric and off-labeled use of drugs, some of them with potentially harmful adverse effects, to the limited number of therapeutics with proven efficacy and safety such as corticosteroids and oxygen (22-26). We could expect that these changes resulted in more favorable economic profiles of novel, modified health care protocols. However, other disturbing trends are developing simultaneously, like the emergence of mutated virus strains with stronger pathogenicity and adverse disease outcomes, as well as exhausting of hospital resources with unapproved therapeutic uses (27, 28). Therefore, future economic research in the field could implement not only more detailed and diverse designs but also historical context of pandemic dynamics.

In conclusion, direct medical treatment costs for patients hospitalized in COVID-19 units of a tertiary care center of developing country during pandemic beginnings were high, representing significant economic burden from the perspective of health insurance payer. We suggest that link between increased costs and unfavorable final outcome should be further explored in the future.

## Data Availability

ll data produced in the present work are contained in the manuscript

## AKNOWLEDGEMENT

The research was partially supported by the grant of The Ministry of Education, Science and Technological Development of the Republic of Serbia No175007. The authors praise the work and dedication of physicians and nurses of COVID-19 hospital units and thanks them for contribution to the patients’ medical documentation on which the large part of the study relied.

## CONFLICT OF INTEREST

Authors declare no conflict of interest.

## ETHICS APPROVAL

The study was approved by the Institutional Ethics Committee and conducted according to principles of Declaration of Helsinki (Decision of the Ethics Committee number 01/20-407, dated April 3^rd^ 2020).

## AUTHOR CONTRIBUTIONS

All authors listed in our manuscript contributed adequately to the development of study design, collection of data, statistical analysis and writing the manuscript of this health economic study to be included as authors, and all of them are listed in the author by-line.

## Notes

### Competing Interest Statement

The authors have declared no competing interest.

### Author Declarations

The study was approved by the Institutional Ethics Committee of University Clinical Centre Kragujevac, Serbia, and conducted according to principles of Declaration of Helsinki (Decision of the Ethics Committee number 01/20-407, dated April 3rd 2020).

## REFERENCES

1. Jin H, Wang H, Li X, et al. Economic burden of COVID-19, China, January-March, 2020: a cost-of-illness study. Bull World Health Organ. 2021; 99(2): 112–124.

2. Nicola M, Alsafi Z, Sohrabi C, et al. The socio-economic implications of the coronavirus pandemic (COVID-19): a review. Int J Surg. 2020; 78: 185–193.

3. Colenda CC, Applegate WB, Reifler BV, et al. COVID-19: financial stress test for academic medical centers. Acad Med. 2020; 95(8): 1143–1145.

4. Harter M, Bremer D, Scherer M, et al. Impact of COVID-19-pandemic on clinical care, work flows and staff at a university hospital: results of an interview-study at the UKE. Gesundheitswesen. 2020; 82(8-09): 676–681.

5. Best MJ, Aziz KT, McFarland EG, et al. Economic implications of decreased elective orthopaedic and musculoskeletal surgery volume during the coronavirus disease 2019 pandemic. Int Orthop. 2020; 44(11): 2221–2228.

6. Shin JH, Takada D, Morishita T, et al. Economic impact of the first wave of the COVID-19 pandemic on acute care hospitals in Japan. PLoS One. 2020; 15(12): e0244852.

7. Khan AA, AlRuthia Y, Balkhi B, et al. Survival and estimation of direct medical costs of hospitalized COVID-19 patients in the Kingdom of Saudi Arabia. Int J Environ Res Public Health. 2020; 17(20): 7458.

8. Cleary SM, Wilkinson T, Tamandjou Tchuem CR, et al. Cost-effectiveness of intensive care for hospitalized COVID-19 patients: experience from South Africa. BMC Health Serv Res. 2021; 21(1): 82.

9. Di Fusco M, Shea KM, Lin J, et al. Health outcomes and economic burden of hospitalized COVID-19 patients in the United States. J Med Econ. 2021; 24(1): 308–317.

10. Lu QB, Jiang WL, Zhang X, et al. Comorbidities for fatal outcome among the COVID-19 patients: A hospital-based case-control study. J Infect. 2020; S0163-4453(20)30507-7.

11. Zhou F, Yu T, Du R, et al. Clinical course and risk factors for mortality of adult in patients with COVID-19 in Wuhan, China: a retrospective cohort study. Lancet. 2020; 395(10229): 1054–1062.

12. World Health Organization (WHO). Global COVID-19 clinical platform. Rapid core case report form (CRF). WHO/2019-nCoV/Clinical_CRF/2020.4. Geneva: World Health Organization, 2020 [cited 2021 May 2]. Available from: https://apps.who.int/iris/rest/bitstreams/1287200/retrieve.

13. World Health Organization (WHO). Clinical management of COVID-19: interim guidance, 27 May 2020. Geneva: World Health Organization, 2020 [cited 2021 May 31]. Available from: https://apps.who.int/iris/handle/10665/332196.

14. World Health Organization (WHO). WHO R&D Blueprint novel Coronavirus COVID-19 Therapeutic trial synopsis. Geneva: World Health Organization, 2020 [cited 2021 May 2]. Available from: https://www.who.int/publications/i/item/covid-19-therapeutic-trial-synopsis.

15. World Health Organization (WHO). ICD-10 Version: 2019. Geneva: World Health Organization, 2019 [cited 2021 May 2]. Available from: https://icd.who.int/browse10/2019/en.

16. Charlson ME, Pompei P, Ales KL, et al. A new method of classifying prognostic comorbidity in longitudinal studies: development and validation. J Chronic Dis. 1987; 40(5): 373–83.

17. World Bank. GDP per capita, PPP (current international $) for 2019. Washington: The World bank, 2019 [cited 2021 May 2]. Available from: https://data.worldbank.org/indicator/NY.GDP.PCAP.PP.CD?most_recent_value_desc=true.

18. United Nations. World Economic Situation Prospects. Statistical annex. New York: United Nations, 2020 [cited 2021 May 2]. Available from: http://www.un.org/development/desa/dpad/wp-content/uploads/sites/45/WESP2020_Annex.pdf.

19. Squire MM, Munsamy M, Lin G, et al. Modeling hospital energy and economic costs for COVID-19 infection control interventions. Energy Build. 2021: 110948. 23.

20. Ignjatovic Ristic D, Hinic D, Bankovic D, et al. Levels of stress and resilience related to the COVID-19 pandemic among academic medical staff in Serbia. Psychiatry Clin Neurosci. 2020; 74(11): 604–605.

21. Nguyen NT Chinn J, Nahmias J, et al. Outcomes and mortality among adults hospitalized with COVID-19 at US medical centers. JAMA Netw Open. 2021; 4: e210417.

22. Milovanovic DR, Jankovic SM, Ruzic-Zecevic D, et al. Treatment of coronavirus disease (COVID-19). Medicinski Casopis 2020; 54(1): 23–43.

23. Jankovic SM. Antiviral therapy of COVID-19. Scripta Medica. 2020; 51(3): 131–33.

24. WHO Solidarity Trial Consortium, Pan H, Peto R, Henao-Restrepo AM, et al. Repurposed antiviral drugs for Covid-19 - Interim WHO Solidarity Trial Results. N Engl J Med. 2021; 384(6): 497–511.

25. RECOVERY Collaborative Group, Horby P, Lim WS, Emberson JR, et al. Dexamethasone in hospitalized patients with Covid-19. N Engl J Med. 2021; 384(8): 693–704.

26. Chalmers JD, Crichton ML, Goeminne PC, et al. Management of hospitalised adults with coronavirus disease 2019 (COVID-19): a European Respiratory Society living guideline. Eur Respir J. 2021; 57(4): 2100048.

27. Vernaz N, Agoritsas T, Calmy A, et al. Early experimental COVID-19 therapies: associations with length of hospital stay, mortality and related costs. Swiss Med Wkly. 2020; 150: w20446.

28. Centers for Disease Control and Prevention. Science brief: emerging SARS-CoV-2 variants. Updated Jan. 28, 2021. Atalanta: Centers for Disease Control and Prevention 2021 [cited 2021 May 2]. Availabe from: https://www.cdc.gov/coronavirus/2019-ncov/science/science-briefs/scientific-brief-emerging-variants.html.

